# Temporal dynamics of human respiratory and gut microbiomes during the course of COVID-19 in adults

**DOI:** 10.1101/2020.07.21.20158758

**Authors:** Rong Xu, Renfei Lu, Tao Zhang, Qunfu Wu, Weihua Cai, Xudong Han, Zhenzhou Wan, Xia Jin, Zhigang Zhang, Chiyu Zhang

**Author notes:** Correspondence: Chiyu Zhang, Shanghai Public Health Clinical Center, Fudan University, Shanghai 201508, China., Or Zhigang Zhang, State Key Laboratory for Conservation and Utilization of Bio-Resources in Yunnan, School of Life Sciences, Yunnan University, No. 2 North Cuihu Road, Kunming, Yunnan 650091, China., Or Xia Jin, Shanghai Public Health Clinical Center, Fudan University, Shanghai 201508, China. These authors contributed equally to this work (Rong Xu, Renfei Lu, Tao Zhang, Qunfu Wu).

## Abstract

SARS-CoV-2 infects multiple organs including the respiratory tract and gut. Whether regional microbiomes are disturbed significantly to affect the disease progression of COVID-19 is largely unknown. To address this question, we performed cross-sectional and longitudinal analyses of throat and anal swabs from 35 COVID-19 adults and 15 controls by 16S rRNA gene sequencing. The results allowed a partitioning of patients into 3-4 categories (I-IV) with distinct microbial community types in both sites. Lower-diversity community types often appeared in the early phase of COVID-19, and synchronous fast restoration of both the respiratory and gut microbiomes from early dysbiosis towards late near-normal was observed in 6/8 mild COVID-19 adult patients despite they had a relatively slow clinical recovery. The synchronous shift of the community types was associated with significantly positive bacterial interactions between the respiratory tract and gut, possibly along the airway-gut axis. These findings reveal previously unknown interactions between respiratory and gut microbiomes, and suggest that modulations of regional microbiota might help to improve the recovery from COVID-19 in adult patients.

## Introduction

COVID-19, a severe respiratory disease caused by a novel virus SARS-CoV-2^1,2^, has led to a devastating global pandemic. It typically presents as asymptomatic infection and mild respiratory symptoms, but in those older than 60 years or having comorbidities, COVID-19 can develop into severe pneumonia and cause death^3,4^. The biological mechanisms behind the varied clinical presentation of disease are not fully understood. Because the microbiota plays a major role in the maintenance of human health by shaping the immune system and maintaining homeostasis^5^, and the microbial composition in the respiratory tract and the gut have been linked to the occurrence and severity of various respiratory viral infections, and affects subsequent respiratory health^6,7^, such as increased airway susceptibility to infection by other RVs and/or colonization of pathogenic bacteria^8-10^, it is reasonable to posit that the new respiratory infection COVID-19 may also interact with microbiota.

Recent studies show that SARS-CoV-2 infects human gut enterocytes and causes diarrhea^11,12^, and COVID-19 patients have altered gut microbiota featuring an enrichment of opportunistic pathogens and a depletion of beneficial bacteria^13,14^. However, the effect of COVID-19 on respiratory microbiome has never been evaluated. Although one study reported persistent alterations in the gut microbiota using longitudinal stool samples collected during COVID-19 patients’ hospitalization^13^, but no study has been performed to simultaneously investigate the temporal dynamics of both the respiratory and gut microbiota at various stages of disease. In this study, we investigated the dynamics of both the respiratory and gut microbiomes in a cohort of COVID-19 patients and controls, and presented a consistent pattern of changes in these two organs from early dysbiosis towards late incomplete restoration in the majority of COVID-19 adults.

## Results Study cohort

The study subjects included 35 adult COVID-19 patients from 17 to 68 years of age, 15 healthy adults, and 10 non-COVID-19 patients with other diseases. All COVID-19 patients (except patient p09) had mild clinical symptoms. A total of 146 specimens including 37 pairs of both throat and anal swabs were collected from COVID-19 patients (Supplementary Fig. S1). High-throughput sequencing of the V4-region of bacterial 16S rRNA gene was performed for all samples (See Supplementary Methods).

## Respiratory microbiome dynamics in COVID-19

The 16S-rRNA gene sequences of all throat swabs were resolved into 3,126 amplicon sequence variants (ASVs) representing 17 known phyla including 209 known genera (Supplementary Table S1). Six throat microbial community types (or clusters) were identified using the Dirichlet Multinomial Mixtures (DMM) modelling based on lowest Laplace approximation (Fig. 1a) and visualized by Nonmetric Multidimensional Scaling (NMDS) based on Bray-Curtis distance (Fig. 1b). Healthy adults (H) and non-COVID-19 patients (NP) formed independent clusters. The vast majority of the specimens of COVID-19 patients were divided into four community types, called I-IV, and 6 specimens were included in the NP type (Fig. 1a). All COVID-19-related community types, as well as the NP type, were significantly separated from the H type. Community types III and IV were not only significantly separated from the types I and II, but also from each other (Fig. 1b). Consistently, significantly lower richness and evenness were observed in community types III and IV, compared with the H type (Fig. 1c).

**Figure 1.**
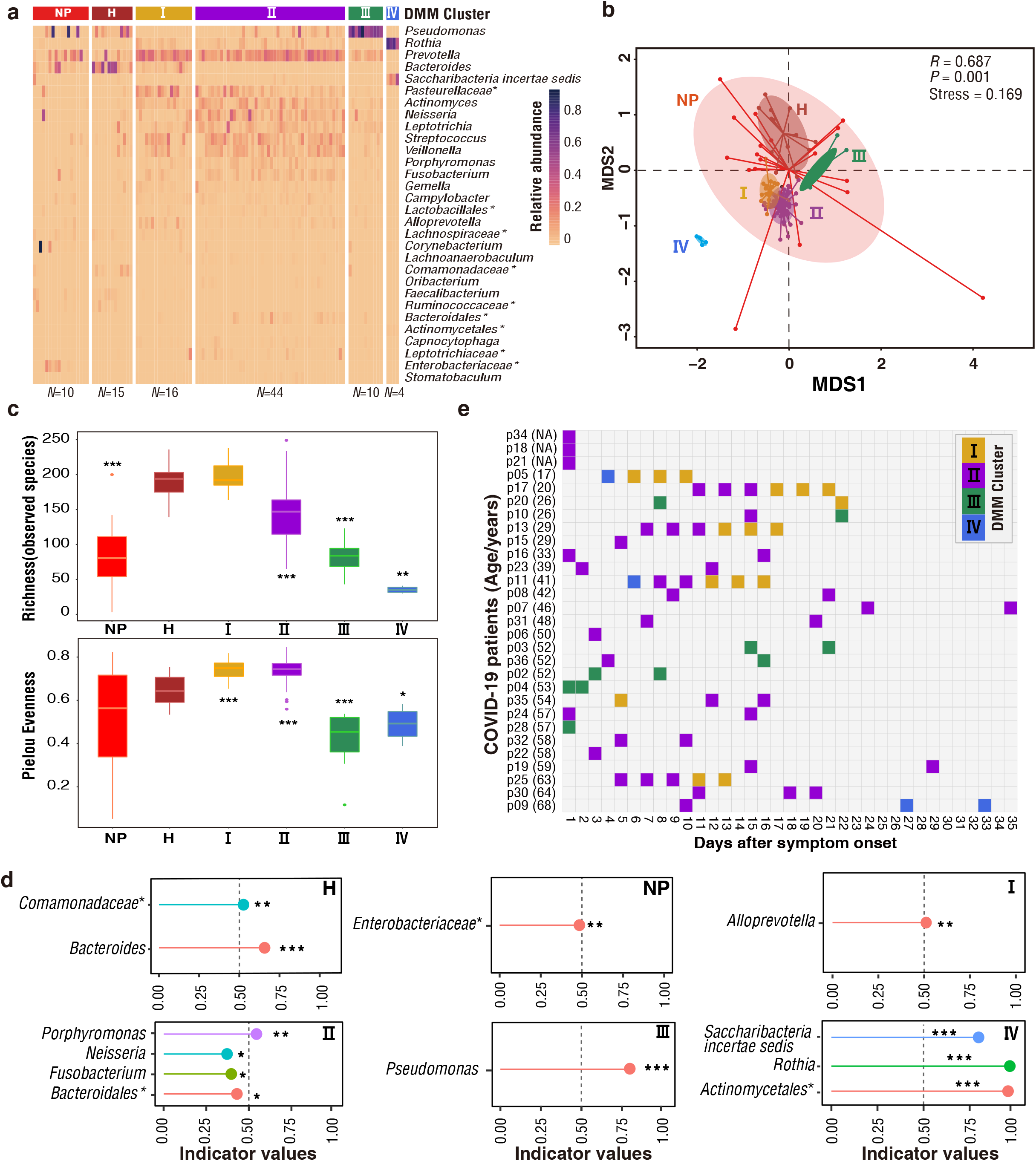
DMM clustering of 16S rRNA gene sequencing data of throat microbiota (*N* = 112). Dirichlet multinomial mixtures (DMM) modelling was applied to 16S rRNA gene sequencing. The entire dataset formed six distinct clusters based on lowest Laplace approximation. Bacterial taxa marked by the stars represent unclassified bacteria genera. a. Heat map showing the relative abundance of the 30 most dominant bacterial genera per DMM cluster. The stars represent unclassified genera. NP, enriched in Non-COVID-19 patients. H, enriched in Healthy individuals. I-IV enriched in COVID-19 patients. b. Nonmetric multidimensional scaling (NMDS) visualization of DMM clusters using Bray-Curtis distance of throat bacterial genera. The ANOSIM statistic R closer to 1 with < 0.05 P value suggest significant separation of microbial community structures. The stress value that was lower than 0.2 provides a good representation in reduced dimensions. c. Box plots showing the alpha diversity (richness and evenness) per each DMM cluster. d. Indicators of airway microbial community types (DMM clusters) identified from top 30 genus contributing to throat microbial community typing (DMM clustering) in a. * P <0.05, ** P <0.01, and *** P < 0.001. e.Dynamic shift of four throat microbial community types (DMM clusters) in different COVID-19 stages. Empty boxes indicate samples were unavailable in COVID-19 patients. Ages (years) were shown in parenthesis. NA, unavailable.

To more directly demonstrate that the variation of throat microbial composition is an indicator of COVID-19 disease stages, the community type-specific indicator taxa were identified based on the top 30 microbial genera (Fig. 1d). The type H was characterized by bacterial genus *Bacteroides* (predominant taxa in the lung of healthy individuals) and unclassified *Comamonadaceae*, whereas the NP type was marked by pro-inflammatory *Enterobacteriaceae* members. In contrast, the indicator bacteria of four COVID-19-related community types were *Alloprevotella* in I, *Porphyromonas, Neisseria, Fusobacterium* and unclassified *Bacteroidales* in II, *Pseudomonas* in III, and *Saccharibacteria incertae sedis, Rothia* and unclassified *Actinomycetales* in IV (Fig. 1d). It is clear that the indicators in the types III and IV belong to putative pathogenic (e.g. *Pseudomonas*) and opportunistic pathogenic bacteria (e.g. *Rothia*). Community type I contained *Alloprevotella* genus, as well as abundant *Bacteroides* and *Prevotella* that typically present in the H type (Fig.1a). Pathogenic bacteria *Pseudomonas, Saccharibacteria incertae sedis* and *Rothia* that are associated with pneumonia and various human diseases were significantly enriched in community types III and IV, reflecting an impaired microbiome (dysbiosis) in the respiratory tract^15-17^. Apart from beneficial commensals (e.g. *Bacteroidales*), some opportunistic pathogenic bacteria such as *Porphyromonas, Fusobacterium*, and *Neisseria* that typically exist in the nasopharynx, and are associated with pneumonia or chronic periodontitis^18-20^, were over represented in type II, possibly reflecting an intermediate state between the normal and dysbiosis in the respiratory microbiota (Supplementary Table S1 and Fig. S2). According to indicator characteristics, community types I to IV reflected a progressive imbalance of the respiratory microbiome.

Longitudinal analysis showed that lower-diversity community types often appeared in early specimens (Fig. 1e), and throat microbiota was dominated by few putative pathogenic and opportunistic bacteria at the early phase of COVID-19 (Supplementary Fig. S2). Prominent microbiome community type shifts from early lower-diversity community types (NP, IV or II) towards later higher-diversity types (II or I) were observed in 9/24 COVID-19 adults who had specimens at two or more time points. For example, an obvious throat microbiome progression from type (IV or II) in early specimens to type I in late specimens was observed in five patients (p17, p25, p13, p11 and p05) with 4 or more consecutive specimens (Fig. 1e). Accompanied with the restoration of throat microbiota, beneficial commensals appeared to occupy the niches, and the bacterial diversity increased accordingly (Supplementary Fig. S2). However, a reversed pattern was observed in four patients who had microbiome composition shift from early higher-diversity types (I or II) to later lower-diversity type (II-IV), implying a worsening of the throat microbiome. For example, in the only severe case (p09), the community type II on day 10 was shifted to type IV on day 27, and sustained to at least day 33 after symptom onset (Fig. 1e), and pathogenic bacteria *Saccharibacteria incertae sedis* and *Rothia* were significantly enriched at late stage (Supplementary Fig. S2). Furthermore, a persistent *Pseudomonas*-dominated throat microbiota (community type III) was observed in three patients (p02, p03 and p04). These results indicate that the change of the respiratory microbiome might be closely associated with the disease progression in COVID-19 patients.

## Gut microbiome dynamics in COVID-19

To expand the scope of this research, a total of 1,940 ASVs were recovered from the 16S-rRNA gene sequences of all anal swabs, representing 13 known phyla including 182 known genera (Supplementary Table S1). The gut microbial communities of COVID-19 patients formed three distinct community types I-III (Fig. 2a-b). The richness and evenness of the gut microbiome decreased from type I to III (Fig. 2c). Indicator analyses showed that type I was primarily characterized by healthy gut bacteria including *Bacteroides* genus and several known butyrate-producing bacteria (e.g. *Faecalibacterium, Roseburia, Blautia*, and *Coprococcus*) and one opportunistic pathogenic bacterium (*Finegoldia*) (Fig. 2d)^21-27^. The indicators of type II mainly contain various pathogenic or opportunistic pathogenic bacteria (e.g. *Neisseria* and *Actinomyces*). In community type III, the gut microbiota was dominated by *Pseudomonas*, implying a severe dysbiosis. We also used the gut community types I-III to reflect the progressive worsening of the microbiome.

**Figure 2.**
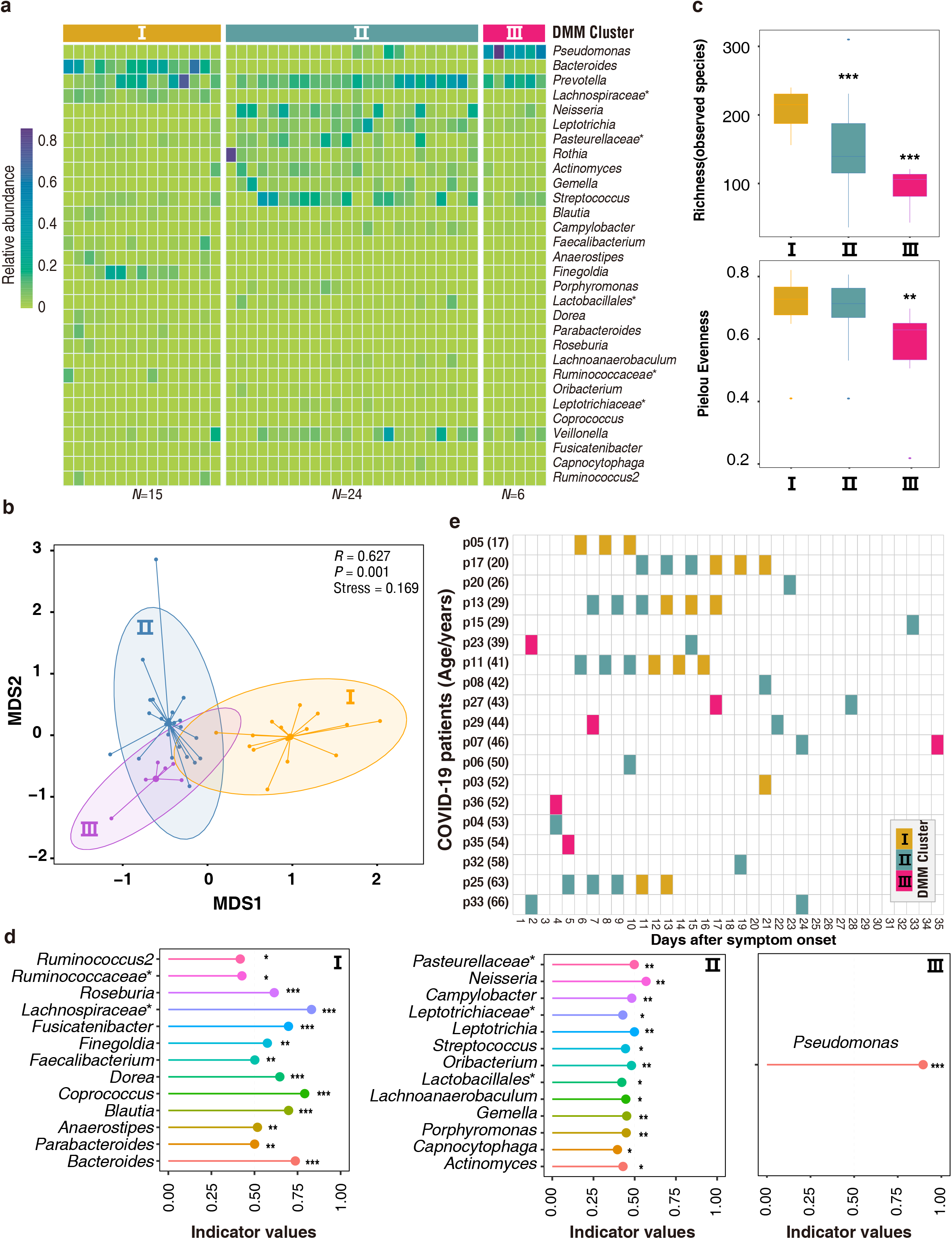
DMM clustering of 16S rRNA gene sequencing data of gut microbiota (*N* = 45). Dirichlet multinomial mixtures (DMM) modelling was applied to 16S rRNA gene sequencing. The entire dataset formed three distinct clusters based on lowest Laplace approximation. All samples were collected from COVID-19 patients. Bacterial taxa marked by the stars represent unclassified bacteria genera. a. Heat map showing the relative abundance of the 30 most dominant bacterial genera per DMM cluster. b. Nonmetric multidimensional scaling (NMDS) visualization of DMM clusters using Bray-Curtis distance of gut bacterial genera. The ANOSIM statistic R closer to 1 with < 0.05 P value suggest significant separation of microbial community structures. The stress value that was lower than 0.2 provides a good representation in reduced dimensions. c. Box plots showing the alpha diversity (richness and evenness) per each DMM cluster. d. Indicators of gut microbial community types (DMM clusters) identified from top 30 genus contributing to gut microbial community typing (DMM clustering) in a. * P <0.05, ** P <0.01, and *** P < 0.001. e. Dynamic shift of gut microbial community types (DMM clusters) in different COVID-19 stages. Empty boxes indicate samples were unavailable in COVID-19 patients. Ages (years) were shown in parenthesis.

A shift of the gut microbiome from the lower-diversity community type (II or III) towards a higher-diversity type (I or II) was observed over time in 8/10 patients who had anal swabs at different time points (Fig. 2e). Accompanied with the shift, a clear trend of increase of bacterial diversity and the relative abundance of beneficial commensals (e.g. *Bacteroides* and *Faecalibacterium*) was observed in the gut microbiota from early to late stages of COVID-19 (Supplementary Fig. S3), indicating the restoration of gut microbiota. Only one patient had a reverse shift from higher-diversity type II to lower-diversity community type III.

## Association between the respiratory and gut microbiomes in COVID-19

Most paired throat and anal swabs showed the same or similar community type levels (Fig. 3a). In particular, the direction of shift over time of the microbiome appeared to match between the throat and the gut in 7/8 patients who had two or more paired specimens at different time points (Fig. 3a). Synchronous improvements of both the respiratory and gut microbiomes from early lower-diversity community type towards late higher-diversity type occurred in six patients (p05, p17, p13, p11, p25 and p29). One patient (p33) showed an improved respiratory microbiome but a stable gut community type on day 24. Only one case (p07) had a worsen gut microbiome from day 24 to day 35 but remained a stable respiratory community type. These results suggested that the microbial community dynamics of either niches may reflect disease development and flora restoration. Because of no available anal specimens, we were unable to assess whether the gut microbiota, like the respiratory microbiota, shifted from higher-diversity type to lower-diversity type over time in this severe case (p09) (Fig. 1e). Except for the duration of COVID-19, the respiratory and gut microbial community divergence seemed not to be significantly correlated with age, gender, antibiotics use, and detection of SARS-CoV-2 RNA (Supplementary Figs. S4 and S5).

**Figure 3.**
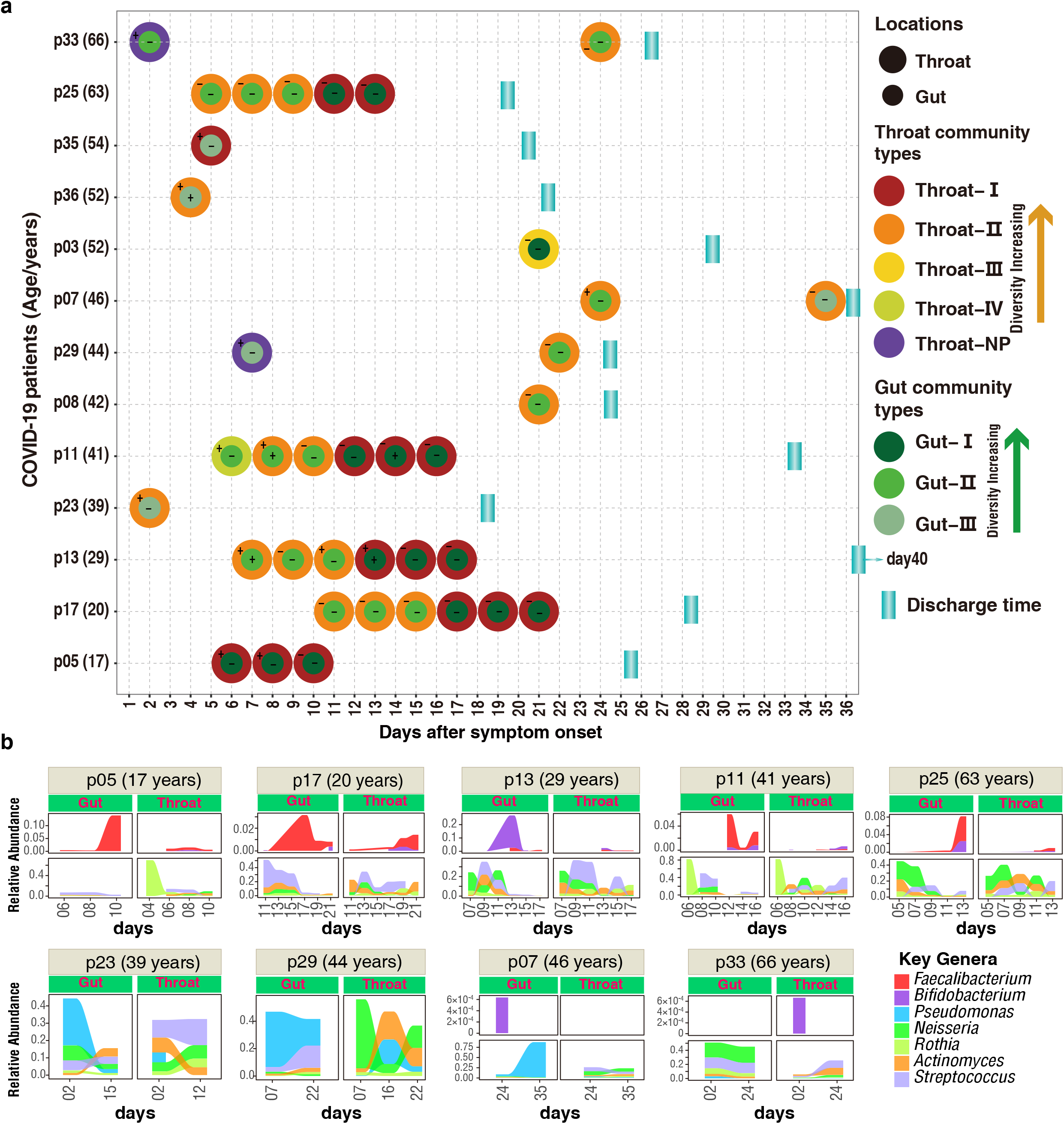
Dynamic change of both DMM clusters (a) and seven key taxa (b) in respiratory tract and gut of patients with mild COVID-19. a, Co-variation dynamics of throat and gut microbial communities of 13 COVID-19 patients. Filled circles indicate the presence of microbial community types. Positive or Negative detections of SARS--CoV-2 in gut or throat are implicated by + or - symbols, respectively. Age (months) of each COVID-19 adult was shown in brackets. b, key taxa of DMM clusters were shown in nine mild COVID-19 adults with at least two time points of sampling. Linked to Fig.1a, Fig.2a, and Supplementary Figs.2-3.

We further assessed the changes of the relative abundance of several representative bacteria, including two probiotics (i.e. *Bifidobacterium* and *Faecalibacterium*), and five pathogenic bacteria (Fig. 3b). The relative abundances of *Bifidobacterium* and *Faecalibacterium* appeared to be negatively correlated with the relative abundance of the five pathogenic bacteria. An obvious decrease in the relative abundance of pathogenic bacteria was accompanied by the restoration of the respiratory and gut microbiome over time in five patients having three or more longitudinal samples (Fig.3b upper panel and Supplementary Fig. S2-S3). Moreover, significantly decreased abundance of *Pseudomonas* was observed in both organs in another two patients (p23 and p29). The relative abundance of *Pseudomonas* increased in the only patient (p07) who had a worsening gut microbiome.

## Bacteria–bacteria co-occurrence networks

There were four indicator bacteria genera (*Porphyromonas, Neisseria*, and *Fusobacterium* in type II and *Pseudomonas* in type III) in the throat microbiomes that had been identified as the indicators of gut microbial community types II and III in COVID-19 patients (Supplementary Fig. S6). Apart from the shared indicators, oropharyngeal pathogenic bacteria *Capnocytophaga* and *Actinomyces* were also identified as indicators of the gut microbial community type II (Figs. 1d and 2d)^28,29^. Because community types II and III often appeared in the early stage of COVID-19 (Figs. 1e and 2e), the appearance of these oropharyngeal bacteria in the gut suggested that a cross-talk between the respiratory and gut microbiomes occurred by frequent bacterial translocation during the early stage.

To further investigate the association between the respiratory and gut microbiomes, we performed co-occurrence network analysis using paired specimens from 13 patients. Significantly positive interactions between the respiratory and gut microbiota were observed (FDR-adjusted P<0.05 and spearman correlation r > 0.7), despite obvious separations of co-occurrence patterns between different community types (Fig. 4). There were four patterns of co-occurrence networks (Throat-IV, Throat-III, II-II and I-I) that had been identified, and only bacteria from the same community types formed significant co-occurrence networks regardless of whether they were in the throat or gut. Under the dysbiosis condition, a few dominant pathogenic bacteria (e.g. *Saccharibacteria incertae sedis, Rothia* and *Pseudomonas*) mediated the bacterial interactions, and drove the oropharyngeal bacterial translocation to the gut possibly due to increased mucosal permeability of the damaged airway and gut (Fig. 4: Throat-III and Throat-IV). Some beneficial bacteria from *Firmicutes* are the primary colonizers to prevent immune-mediated diseases^21^. Accompanied with the increase of bacteria diversity, more gut bacteria started to interact with throat microbiota, and some beneficial commensals (mainly *Firmicutes*) occupied the niches of the throat and gut mucosa to compete with the pathogenic bacteria (Fig. 4: II-II), leading to a possible restoration of the damaged microbiota in both organs. At the late stage of COVID-19, the species from *Firmicutes* and *Bacterioidetes* dominated the bacterial community in both organs (Fig. 4: I-I). In particular, the appearance of various probiotics (e.g. *Bifidobacterium* and *Faecalibacterium*) in both the throat and gut implied further improvement and restoration of the respiratory and gut microbiomes^25,30^.

**Figure 4.**
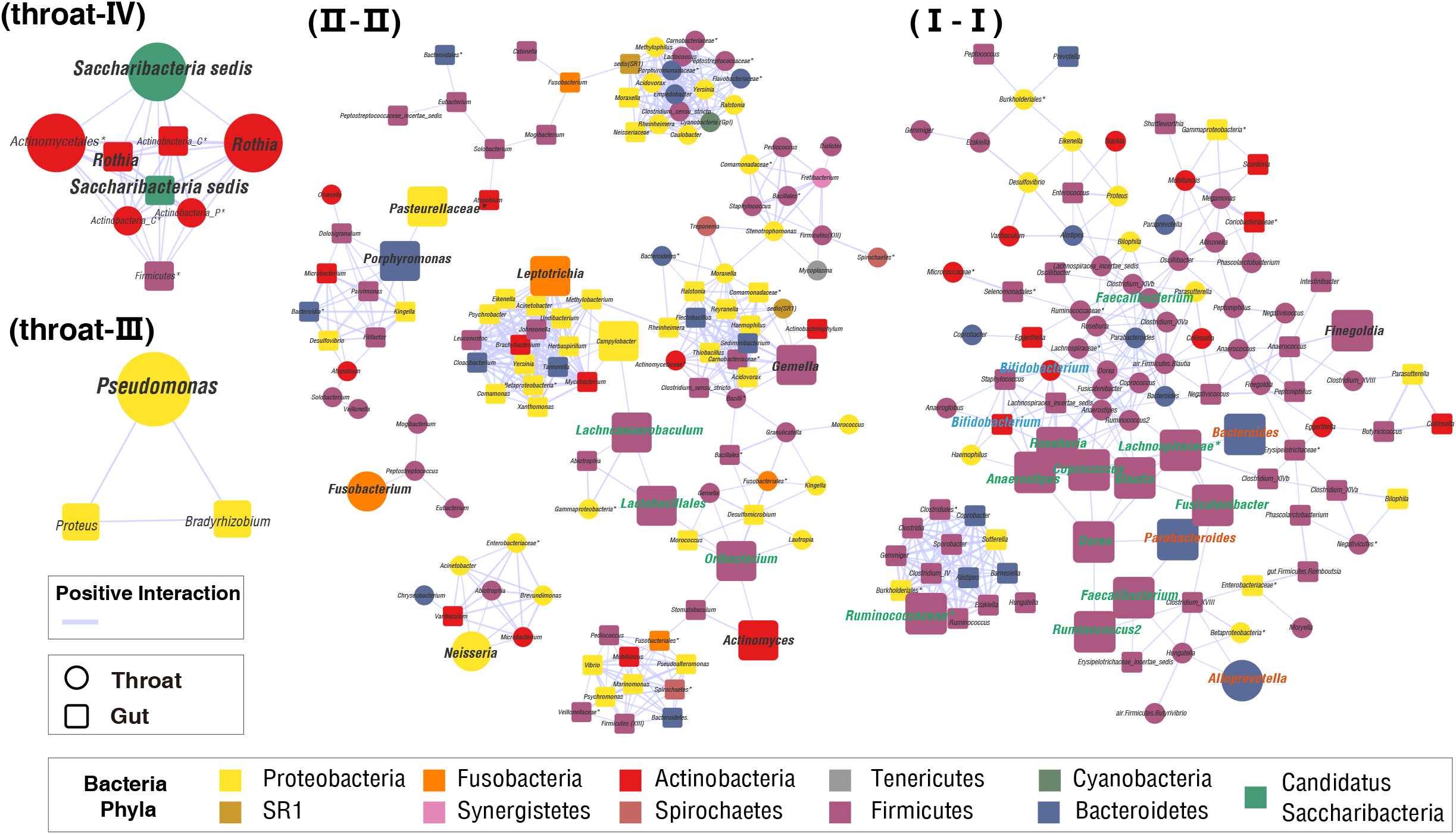
Co-occurrence networks of gut and throat microbiota within 13 COVID-19 patients. Pearson correlation was employed to calculate correlation coefficient (r) between bacterial genus pairs based on their relative abundances. Co-occurred pairs with r > 0.7 under FDR-adjusted P < 0.05 were shown and visualized by Cytoscape version 3.8.0. Negative correlations that were not significant were not shown. Edges were sized based on r values. The stars represented unclassified bacteria genera. Four co-occurrence patterns mediated by throat-IV indicator, throat-III indicator, throat-II & gut-II indicators, and throat-I& gut-I indicators, respectively. The bigger squares or circles were indicators in Figs. 1d and 2d.

## Discussion

Whether SARS-CoV-2 infection alters microbiota to affect COVID-19 disease progression is an important question that needs answers. In this study, we made three major observations. First, the respiratory and gut microbiota compositions of COVID-19 adults can be characterized by four (I-IV) and three (I-III) community types, respectively, and these types reflect different levels of balance between the near-normal microbiota (type I) and dysbiosis (type III/IV). Second, lower-diversity community types III/IV often appears in the early phase of COVID-19, and a consistent pattern of shift from early dysbiosis towards late near-normal coincides with the recovery of both the respiratory and gut microbiomes in most patients with mild COVID-19. Third, the shift of community types is synchronous in the respiratory tract and gut.

SARS-CoV-2 infects cells through ACE2 receptor^2^, which is highly expressed in respiratory and intestinal epithelial cells^31^. SARS-CoV-2 infection can trigger the cytokine storm, cause local pathological damage^32,33^. As an open system with direct contact with environment and the primary site for respiratory infections, the respiratory tract microbiota is more easily affected by SARS-CoV-2 infection, but has not been examined yet. We observed alterations of the respiratory microbiota in COVID-19 adults, and presented data on the dynamic change of the respiratory microbiome composition over time. The respiratory microbiome of the COVID-19 adults was characterized by four bacterial community types I-IV, which reflect the different levels of the normal microbiome to dysbiosis. The community type III dominated by *Pseudomonas* and type IV enriched for other three pathogenic bacteria often appeared in early throat specimens (e.g. first several days after symptom onset), indicating SARS-CoV-2 infection results in a very rapid dysbiosis in respiratory tract. A restoration of the respiratory microbiome from dysbiosis towards near-normal was observed over time in some with mild disease, whereas prolonged or worsening microbiome appeared in a few others including the only one severe case (p09). The inconsistency indicates that the diversity characteristics of the respiratory microbiome had been affected by COVID-19.

Intestinal enterocytes that express ACE2 are also the target of SARS-CoV-2 which further up-regulates the expression of ACE2, leading to a longer viral RNA shedding time in the gut than respiratory tract^11,31^. The baseline microbiome composition with abundant pathogenic bacteria (e.g. *Coprobacillus, Clostridium ramoaum* and *Clostridium hathewayi*) had been associated with the fecal levels of SARS-CoV-2 and COVID-19 severity in a previous study^31^. However, the sampling time was relatively late in that study (about 14 days after symptom onset), therefore unable to determine whether the baseline microbiome status is a consequence of SARS-CoV-2 infection, or a cause of disease severity. We also observed alterations of the gut microbiota during COVID-19 in adults, and found some pathogenic bacteria (e.g. *Streptococcus, Rothia, Veillonella, Actinomyces, Bacteroides* and *Actinomyces*) similar to those reported in the previous observations^13,14^. However,distinct from the previous studies, three community types I-III were identified to characterize the changes of gut microbiome over time, and low-diversity community types II and III often appeared in early specimens, supporting the early effect of SARS-CoV-2 on the microbiome. A fast restoration with the community type shifted from low-diversity type II to high-diversity type I over time was observed in at least 4 patients. Moreover, the *Pseudomonas-*dominated community type III appeared in earlier specimens of patients, and showed a slow improvement towards community II in three patients. Of particular importance is that temporal dynamic changes of the respiratory and gut microbiomes matched with each other extremely well, indicating a close association in microbiota between both body sites, possibly via the “airway-gut axis”^34^.

The reason for the fast dysbiosis in both the respiratory tract and gut of patients with COVID-19 might be that the early-stage inflammation induced by SARS-CoV-2 infection damages the mucosal tissues in airway, lung and gut^7,35^, and results in a fast loss of beneficial commensals, which then augments the colonization and growth of pathogens (Supplementary Fig. S7). The use of empirical antibiotics in some patient during the early stages of the pandemic may exacerbate the dysbiosis in the respiratory tract and gut. Therefore, the microbiome composition with enrichment of pathogenic bacteria (e.g. *Pseudomonas* and *Neisseria*) was observed in both throat and gut microbiomes during the first several days after symptom onsets. Because the respiratory tract is more receptive to both exogenous and indigenous microbes than the gut^7,36^, the dysbiosis of respiratory microbiome appeared to be worse and occurred earlier than that of the gut microbiota, as manifested by lower diversity and richness and more indicators of presumed pathogenic bacteria in the former than in the latter. During this early phase, the damaged respiratory tract mucosa enables some invasive respiratory pathogenic bacteria to be translocated to the gut, worsening the gut bacterial community (Supplementary Fig. S7).

Gut microbiota plays an important role in human health by shaping local immunity and remodeling mucosal tissues^37^. It is relatively more stable and plastic than the respiratory microbiota, and it may affect the latter by cross-talk between these two organs along the airway-gut axis^34,36^, as determined by the significantly positive co-occurrence relationship between the bacteria of the gut and respiratory tract. In spite of longer duration of SARS-CoV-2 shedding in the gut than in the respiratory tract, gut microbiota appeared to have a faster restoration with an increased bacterial diversity and enrichment of beneficial commensals than the respiratory microbiota. The former then promoted the restoration of the latter via bacterial cross-talk, and resulted in a synchronous restoration of both organs (Fig. 3b, Supplementary Figs. S2 and S3).

There are at least three major stages reflecting the restoration or worsening of the microbiome in COVID-19 adults. The restoration of the microbiome seemed to be independent of early microbiome community types. For example, several patients remained microbiome community type II for a long time. Age, gender and antibiotics use seemed not to be linked to restoration of the microbiome (Supplementary Fig. S4-S5), implying potential contributions from other factors such as diet and genetic background.

We noted that *Pseudomonas-*dominated bacterial community type III was difficult to restore towards higher-diversity community types in the respiratory tract. *Pseudomonas* is a well-known pathogenic bacterium, and rarely found in healthy individuals. The identification of early microbiome dysbiosis community type III or IV dominated by *Pseudomonas, Rothia*, and *Saccharibacteria* in COVID-19 patients might imply a need for microbiota-based personalized antibiotics treatment against these specific pathogens. The community type II represents a crucial intermediate stage during the restoration of the microbiome from dysbiosis towards near-normal. It was characterized by *Neisseria, Fusobacterium*, and *Porphyromonas. Fusobacterium. Porphyromonas* are the common commensals in the oropharynx and the gut^18,20^, while *Neisseria* generally presents in the lung. The appearance of lung *Neisseria* in both the respiratory tract and the gut, implying bacteria translocations along the “airway-lung-gut axis”. The bacteria translocations may be the consequence of increased permeability among these organs caused by local inflammation^38^. The *Bifidobacterium* and some butyrate-producing bacteria (e.g. *Faecalibacterium*) can improve the inflammatory conditions and regulate innate immunity by down-regulating ACE2 expression, and activating the corresponding signaling pathways^25,30^. During the restoration of the microbiota, these probiotics started to occupy the ecological niches in the gut and respiratory tract in the type II stage, and governed the microbial communities in both organs by mass replacement of pathogenic bacteria (e.g. *Rothia* and *Neisseria*) in the type I stage (Fig. 3b and Supplementary Fig. S7). However, a progressively worsening in the respiratory and gut microbiome might be associated with severe cases of COVID-19.

In summary, we revealed for the first time the associations between the respiratory and gut microbiota and COVID-19 disease progression, and observed early dysbiosis towards later restoration to near-normal microbiota in a proportion of adults with mild COVID-19. In the absence of specific antiviral drugs and vaccines for COVID-19, our findings have important clinical implications. First, some indicator bacteria (e.g. pathogenic *pseudomonas* and beneficial butyrate-producing bacteria) can be used as crucial biomarkers for clinical treatment decision making and prognostic markers. The measurement of predominant short-chain fatty acid (especially butyrate) concentration in fecal samples will be particularly useful in early clinical diagnosis. Second, apart from the routine treatment efforts (e.g. non-specific antiviral and supportive treatments)^39^, precision intervention and modulation of the gut and respiratory microbiota may open up novel therapeutic alternatives, such as personalized antibiotics therapy to inhibit certain pathogenic bacteria (e.g. *pseudomonas*). Third, COVID-19 tailored probiotics (e.g. *Bifidobacterium* and *Faecalibacterium*), prebiotics (e.g. xylooligosaccharide) treatment, or symbiotic treatments might be applied to modulate the gut and respiratory microbiota to facilitate the recovery of COVID-19 patients. Lastly, fecal microbiota transplantation may be considered as another treatment choice.

## Methods

### Study population

A total of 63 subjects, including 35 laboratory-confirmed COVID-19 patients, 10 SARS-CoV-2 negative patients with various diseases (non-COVID-19) and 15 healthy adults were enrolled in this study. Demographic and clinical characteristics of these patients were provided in Supplementary Table S2 and S3^40^. Specimens including throat swabs and anal swabs were collected from the patients during hospitalization (10-40) at Nantong Third Hospital Affiliated to Nantong University and from healthy adults when they visited the same hospital for physical examination. Sampling was performed using flexible, sterile, dry swabs, which can reach the posterior oropharynx and anus easily (approximately 2 inches) by the professionals at the hospital. At least two throat swabs at different days were available for 32 of 38 COVID-19 patients (Supplementary Fig S1).

The study was approved by Nantong Third Hospital Ethics Committee (EL2020006: 28 February 2020). Written informed consents were obtained from each of the involved individuals. All experiments were performed in accordance with relevant guidelines and regulations.

### Confirmation of COVID-19 patients

COVID-19 was diagnosed in adult patients according to the National Guidelines for Diagnosis and Treatment of COVID-19. The virus RNA was extracted from all samples using a Mag-Bind RNA Extraction Kit (MACCURA, Sichuan, China) according to the manufacturer’s instructions. Then the *ORFlab* and *N* genes of SARS-CoV-2 was detected using a Novel Coronavirus (2019-nCoV) Real Time RT-PCR Kit (Liferiver, Shanghai, China) according to the manufacturer’s instructions. Only the individuals who had at least two consecutive throat swabs been positive for both *ORFlab* and *N* genes of SARS-CoV-2 were defined as COVID-19 patients. All positive specimens of COVID-19 patients were confirmed by Nantong Center for Disease Control and Prevention (CDC) using recommended real-time RT-PCR assay by China CDC.

### 16S rRNA gene sequencing

Bacterial DNA was extracted from the swabs using a QIAamp DNA Microbiome Kit (QIAGEN, Düsseldorf, Germany) according to the manufacturer’s instructions, and eluted with Nuclease-free water and stored at -80°C until use. The V4 hypervariable region (515-806 nt) of the 16S rRNA gene was amplified universal bacterial primers^41^. To pool and sort multiple samples in a single tube of reactions, two rounds of PCR amplifications were performed using a novel triple-index amplicon sequencing strategy as described previously^42^. The first round of the PCR (PCR1) amplification was performed with a reaction mixture containing 8 μL Nuclease-free water, 0.5 μL KOD-Plus-Neo (TOYOBO, Osaka Boseki, Japan), 2.5 μL of 1 μM PCR1 forward primer, 2.5 μL of 1 μM PCR1 reverse primer, and 5 μL DNA template. The products of the PCR1 reactions were verified using a 1.5% agarose gel, purified using Monarch DNA Gel Extraction Kit (New England Biolabs, Ipswich, MA, USA), and quantified by a Qubit® 4.0 Fluorometer (Invitrogen, Carlsbad, CA, USA). Equal amounts of purified PXR1 products were pooled, and subjected to the secondary round of PCR (PCR2) amplification. The PCR2 was performed with a reaction mix containing 21 μL Nuclease-free water, 1 μL KOD-Plus-Neo (TOYOBO, Osaka Boseki, Japan), 5 μL of 1 μM PCR2 forward primer, 5 μL of 1 μM PCR2 reverse primer, and 5 μL pooled PCR1 products. The PCR2 products were verified using a 2% agarose gel, purified using the same Gel Extraction Kit and qualified using the Qubit® 4.0 Fluorometer. The amounts of the specific product bands were further qualified by Agilent 2100 Bioanalyzer (Agilent, Santa Clara, CA, USA). Equal molars of specific products were pooled and purified after mixing with AMPure XP beads (Beckman Coulter, Pasadena, CA, USA) in a ratio of 0.8:1. Purified amplicons were paired-end sequenced (2×250) using Illumina-P250 sequencer.

### Bioinformatic analysis of 16S rRNA gene sequence data

Sequenced forward and reverse reads were merged using USEARCH11 software^43^, then de-multiplexed according to known barcodes using FASTX-Toolkit^44^. After trimming barcode, adapter and primer sequences using USEARCH11, 19,096,003 sequences were retained with an average of 105508 sequences per sample. Samples with sequence <1000 were excluded from the following analysis.

Because traditional OTU (operational taxonomic units) picking based on a 97% sequence similarity threshold may miss subtle and real biological sequence variation^45^, several novel methods such as DADA2^46^ and Deblur^47^ were developed to resolve sequence data into single-sequence variants. Here, the DADA2 was employed to perform quality control, dereplicate, chimeras remove on Qiime2 platform^48^ with default settings except for truncating sequence length to 250bp. Finally, an amplicon sequence variant (ASV) table, equivalent to OTU table, was generated and then spitted into gut ASV table (2348 ASVs) and throat ASV table (4050 ASVs). The taxonomic classification of ASV representative sequences was conducted by using the RDP Naive Bayesian Classifier algorithm^49^ based on the Ribosomal Database project (RDP) 16S rRNA training set (v16) database^50^. To eliminate sequencing bias across all samples, both the gut ASV table and throat ASV table were subsampled at an even depth of 4700 and 3000 sequences per sample, respectively. The ASV coverage of 82.6% (gut) and 77.2% (throat) were sufficient to capture microbial diversity of both sites.

## Identification and characterization of microbial community types

Dirichlet multinomial mixtures (DMM)^51^ is an algorithm that can efficiently cluster samples based on microbial composition, its sensitivity, reliability and accuracy had been confirmed in many microbiome studies^52-54^. DMM clustering were conducted with bacterial genus abundance from throat and gut microbiota using the command “get.communitytype” introduced by v1.44.1 of mothur^55^. The appropriate microbial community type numbers (DMM clusters) were determined based on the lowest Laplace approximation index. According to sample counts per cluster, the fisher exact test was applied to discover significant associations between each cluster and host conditions (such as healthy controls, COVID-19 patients, and Non-COVID-19 patients) under *P* values that are below 0.05 adjusted by the False Discovery Rate (FDR). Conjugated with the Analysis of Similarities (ANOSIM), the reliability of DMM clustering was further validated and then visualized by the Non-metric multidimensional scaling (NMDS) based on the Bray-Curtis distance under bacterial genus level. “The ANOSIM statistic “R” compares the mean of ranked dissimilarities between groups to the mean of ranked dissimilarities within groups. An R value close to “1.0” indicates dissimilarity between groups, whereas an R value close to “0” indicates an even distribution of high and low ranks within and between groups”. The ANOSIM statistic R always ranges between -1 to 1. The positive R values closer to 1 suggest more similarity within sites than between sites, and that close to 0 represent no difference between sites or within sites^56^. ANOSIM p values that are lower than 0.05 imply a higher similarity within sites. Richness (Observed OTUs/ASVs) and Pielou’s or Species evenness for each community type were calculated for estimating the difference of alpha diversity. The analyses of alpha diversity, NMDS and ANOSIM were performed using R package “vegan” v2.5-6. Dynamic change of community types was showed according to collected dates of specimens with R package ‘P heatmap’. For association between community types and potential confounding factors such as sex, age, virus existence and antibiotic use, the fisher exact test based on sample count was performed and the association with FDR-corrected p value <0.05 was considered significant.

## Indicator analysis in throat and gut community types

According to the definition given by the United Nations Environment Programme (1996), the indicator species are a group of species whose status provides information on the overall condition of the ecosystem and of other species in that ecosystem, reflecting the quality and changes in environmental conditions as well as aspects of community composition. To obtain the reliable indicator genus that is specific to each community type, we performed the Indicator Species Analysis using the indicspecies package (ver.1.7.8) ^57^ in R platform with top 30 genus contributing to DMM clustering in both throat (accounting for 66% cumulative difference) and gut (68% cumulative difference). Dynamic changes of indicator genera corresponding to each throat community type were showed in all COVID-19 patients using the pheatmap package in R and only gut indicator genera with indicator values that were above 0.05 were presented in the patients.

## Co-occurrence network analysis of a crosstalk between throat and gut microbiota

Based on microbial genus abundances of both throat and gut samples collected from 13 COVID-19 patients at the same time point, we calculated the Pearson Correlation Coefficient (Pearson’s r) among the throat & gut microbial genera. The Pearson’s r with *P* values that were below 0.05 after the FDR adjustment were considered significant correlations. Co-occurrence network of significantly correlated microbial genus pairs was visualized using Cytoscape v3.8.0^58^.

## Data Availability

The raw data of 16S rRNA gene sequences are available at NCBI Sequence Read Archive (SRA) (https://www.ncbi.nlm.nih.gov/sra/) at BioProject ID PRJNA639286.

## Supplemental information

Supplemental information includes supplemental Experimental Procedures, six figures, and three tables and can be found with this article.

## Acknowledgements

We thank Miss Yingying Ma at Shanghai Public Health Clinical Center, Fudan University, and Mr. Kai Liu and Mrs Xiuming Wu at Institut Pasteur of Shanghai, Chinese Academy of Sciences for their technical support. This work was supported by grants from the National Key Research and Development Program of China (2017Z×10103009-002, 2019YFC1200603 and 2018YFC2000500), Special fund for COVID-19 diagnosis and treatment of Nantong Science and Technology Bureau (SFCDT3-2), the Second Tibetan Plateau Scientific Expedition and Research (STEP) program (2019QZKK0503), the Key Research Program of the Chinese Academy of Sciences (FZDSW-219), and the Chinese National Natural Science Foundation (31970571).

## Author contributions

C.Z. conceived the study idea. C.Z. and Z.Z. designed and supervised the study. R.L., W.C. and X.H. collected clinical samples and data. R.X. and R.L. performed the experiments. T.Z. and Q.W. processed and analyzed the raw sequencing data. R.X., R.L. and Z.W. analyzed the clinical data. Z.Z. and R.X. generated the figures. C.Z., Z.Z. and X.J. interpreted the data. C.Z., Z.Z., R.X., and T.Z. wrote the first draft of the manuscript. X.J. contributed to critical revision. All authors contributed to the final manuscript.

## Competing interests

The authors have not conflict of interests.

## Supplementary materials

**Supplementary table S1. Throat and gut microbial abundances (phyla and genera)**.

**Supplementary table S2. Clinical index of COVID-19 patients in this study**.

**Supplementary table S3. Demographic information of COVID-19 patients in this study**.

**Supplementary figure S1. COIVD-19 patient admission and discharge time as well as the point of detection of SARS-CoV-2**. a. the hospitalization of p13 was 40 days. b. the information of these patients was unavailable. DAY 1 was the day of symptom onset.

**Supplementary figure S2. Time-scale changes of indicators of throat microbial community types**. Color sectors represent relative abundance of indicators in different COVID-19 stages. Linked to **Figure 1**.

**Supplementary Figure S3. Time-scale changes of indicators of gut microbial community types**. Color sectors represent relative abundance of indicators in different COVID-19 stages. Linked to **Figure 2**.

**Supplementary Figure S4. Enrichment analysis of impact factors on throat microbial community typing**. a) Sex, b) Age, c) Virus detection, and d) Antibiotic uses. Enrichment analysis was performed by using the Fisher’s exact test under FDR-adjusted P < 0.05. Sample numbers were shown on the bar. Only COVID-19 patients were used for this analysis.

**Supplementary Figure S5. Enrichment analysis of impact factors on gut microbial community typing**. a) Sex, b) Age, c) Virus detection, and d) Antibiotic uses. Enrichment analysis was performed by using the Fisher’s exact test under FDR-adjusted P < 0.05. No significant enrichment was observed. Sample numbers were shown on the bar.

**Supplementary Figure S6. Comparisons of indicator genera between throat and gut microbial clusters**.

**Supplementary Figure S7. Putative restoration model of the respiratory and gut microbiomes over time in adults with mild COVID-19**. SARS-CoV-2 infection resulted in a fast dysbiosis in both the respiratory tract and gut at the very early phase of the disease. A fast restoration of both the respiratory and gut microbiomes from early dysbiosis towards late near-normal status was observed in most adults with mild COVID-19 albeit they seemed to have a relatively slow clinical recovery. This model reflects the major microbiome dynamic change in most adults with mild COVID-19 but not all features in all patients, especially in those with severe disease.

## Notes

### Competing Interest Statement

The authors have declared no competing interest.

### Funding Statement

This work was supported by grants from the National Key Research and Development Program of China (2017ZX10103009-002, 2019YFC1200603 and 2018YFC2000500), Special fund for COVID-19 diagnosis and treatment of Nantong Science and Technology Bureau (SFCDT3-2), the Second Tibetan Plateau Scientific Expedition and Research (STEP) program (2019QZKK0503), the Key Research Program of the Chinese Academy of Sciences (FZDSW-219), and the Chinese National Natural Science Foundation (31970571).

